# “Sharing my story has been part of my healing process”: how Native women survivors heal from IPV

**DOI:** 10.1101/2022.12.06.22283188

**Authors:** Susanna Y Park

## Abstract

Four out of five Native women experience intimate partner violence (IPV) in their lifetime. Culture, Native identity, and other socioeconomic factors influence how survivors experience seeking support and healing. High rates of IPV among Native women indicate an urgent public health need to address the long-term implications of being a survivor. This study explores how Native women in California seek healing as survivors of violence through stories shared in Community Circles and Oral Histories supported by collaborations with local Indigenous community organizations in Southern California. Seeking support and healing for Native women survivors of IPV are layered with their identities as Indigenous women, historical oppression, and its resulting intergenerational traumas. Healing from the violence has cultural implications, amplifying the need for community-based systems of support that are Indigenous-centered. For AIAN women survivors of IPV, healing transcends beyond their selves, extending to their families, tribal communities, and future generations. Culturally-rooted systems of support that cultivate Indigenous values of connection and healing are needed.

## Introduction

American Indian/Alaska Native (AIAN) women experience high rates of violence [1–5], with intimate partner violence (IPV) being the most common type of violence they experience [2,6–8]. Nearly four out of five Native women experience IPV in their lifetime. Though data on violence against Indigenous women living on Native American reservation lands and urban settings are sparse, existing studies indicate that AIAN women are at a significantly higher risk of experiencing stalking (48.4%) and physical violence (55.5%) by an intimate partner in their lifetime than non-Hispanic White-only women [4,6,7]. In addition, AIAN women are significantly more likely to experience psychological aggression (66.4%) by an intimate partner in their lifetime in the past year [8]. Over two out of three AIAN women will experience sexual assault in their lifetime [3]. Qualitative research on the experiences of violence among AIAN women suggests that the level of physical violence is more severe for Native women compared to women belonging to other ethnic groups. Over 90% reported being physically assaulted and 25% reported that their perpetrator used a weapon [6]. To compare with White women, 71% reported physical assault and 9% reported the use of a weapon [6,7].

Public health research thus far has focused on gathering prevalence data and identifying protective and risk factors to better tailor interventions. However, not much is known on the support-seeking behaviors among AIAN women; culturally rooted research on how AIAN women seek support as survivors is even more sparse. Programs addressing violence vary in scope: community education, health professionals training on treating victims of violence, and telephone helplines. Programs that are specifically created for Native communities are infrequent. IPV is linked to negative health outcomes, and can severely impact a woman’s physical, mental, and emotional health [4,9]. It is also an issue that continues throughout the lifespan, affecting women in all stages of life and occurring multiple times [10]. There is a clear need to address such impacts of IPV for AIAN women who are at significant risk of experiencing IPV.

Studies examining support-seeking behavior of women who experience IPV suggest that they are more likely to seek informal support (e.g. friends and family) than seek formal support (e.g. police or medical) [11–13]. If women perceive the violence as justified or engage in self-blaming behavior, they may be less likely to seek support [11]. Informal support may often lead to eventually seeking formal support. For instance, domestic violence survivors who sought informal support also decided to seek formal help if the family or friend they disclosed to had experienced or had knowledge of domestic violence [14]. For AIAN women, informal support is often protective if the family or social network is tight-knit, supportive, and affirming. However, if there is a risk of breaching confidentiality and lack of accountability [15], AIAN women are less likely to access informal networks for support and may seek formal support or not seek support at all [16].

This study aims to contribute to current understandings of how Native women survivors of IPV seek support through the lived experiences of survivors in California and learning about their journeys to find healing.

## Methods

### Conceptual framework

This study specifically draws from Burnette and Figley’s (2017) ecosystemic framework on historical oppression, resilience, and resistance. Historical oppression as a societal level risk factor for violence that permeates through other levels of the framework recognizes that IPV victimization transcends beyond individual-level impacts. The interconnectedness across individual, family and relational, community and cultural, and societal levels is not only congruent to the Indigenous worldview but allows for a nuanced exploration of how survivors seek support [17–19].

The individual exists as part of an ecosystem that cannot be understood without the other levels [20]. Individual experiences with seeking support relate to family and relational factors, such as their parents, extended family, or friends. In most IPV cases, individuals tend to seek help from their informal networks [11–13]. However, individuals may further pursue seeking support from a formal network if their friends and family support them seeking professional help [15]. On the other hand, individuals may experience shaming from their informal network and choose to seek support from a formal network but advocate for themselves independently to see effect [21].

How attitudes and perceptions towards violence manifest at these levels are influenced by community and cultural factors, such as family values, community rules, or tribal traditions. In Native communities, a “tight-knit” community may serve as both a positive and negative factor to seeking help. Tight-knit communities can be a source of support and encourage the individual to seek help. On the other hand, the community may be so tight-knit that the individual may choose not to seek help in fear of their confidentiality being breached (Fiolet, Tarzia, Hameed, & Hegarty, 2019). Finally, societal factors like historical oppression play a critical role in informing the structures in which formal services can be accessed by survivors and the perversion of Indigenous values through colonial patriarchy that then permeates across multiple levels. For instance, the loss of “bodily sovereignty” not only through colonialism, but colonialism manifested through acts of violence against Indigenous women [22].

We expect to interpret the findings from this research using this framework. In doing so, the narratives of survivors are to be understood as existing within an ecosystem of interconnected levels. Survivor stories of seeking support and healing are in relative to their identity as Indigenous women, their culture and tribal communities, and their loved ones.

### Study methods

Wilson (2008) uses the term “strategies of inquiry” to describe a process in Indigenous research in which a general direction for the research is strategized, allowing for flexibility and adaptation along the way. Rather than being confined to a list of methodological approaches, he uses a combination of methods that reflect Indigenous research values of “learning by watching and doing” [19]. In the same vein, this project drew from various qualitative methods to reflect Indigenous ways of knowing, relationship building, and relational accountability.

For a year, I engaged in participant observation through feminist critical ethnography, an ethnographic method in which structural inequities through the lenses of race, disability, gender, etc. are assumed to be divided hierarchically, therefore influencing how society functions and influences cultural groups [23]. The feminist lens centers the voices of AIAN women survivors, bound within the context of colonialism and patriarchy as oppressive structural constructions that shape the experiences of women. Contrary to realist ethnography where the researcher aims to remain value neutral and offers an interpretation of observations from the field, critical ethnography values the relationship built with the participants and intentionally recognizes the non-immunity to bias and researcher subjectivity [23,24]. Using feminist critical ethnography allowed me to observe and participate in such spaces with the assumption that people’s lived experiences are affected by oppressive structures of power and privilege [17,24,25].

Participant observation during this stage included attending various Indigenous community events, such as talking circles, pow wows, advocacy efforts for violence against Native women, and educational sessions (e.g., webinars) by Indigenous scholars. A journal was actively kept that reflected on such events, detailing my positionality relative to the event participants and cultural observations. In doing so, I fostered relationships with AIAN community members who work in service sectors, such as family housing, therapy, and medical care.

As I connected with community members and spoke with other Indigenous people about violence against Native women, community members facilitated my ability to connect with key individuals. Key community leaders in California were identified at two AIAN-centered organizations that provide mental and behavioral health services to Native people: Indigenous Circle of Wellness and Seven Generations Child and Family Services. I inquired with the community leaders if they would be willing to read a research brief to potentially collaborate on this research. Partnership was first established with the Indigenous Circle of Wellness, and it was the primary organization that supported the research direction. I met frequently with the Indigenous Circle of Wellness to discuss research methods, accountability, and information dissemination. The Indigenous Circle of Wellness and Seven Generations Child and Family Services were fundamental to the design and recruitment of participants for the study and mirrored the values of Indigenous methodologies that center the voices of participants in the study, are community-centered, and recognize that there are multiples ways of knowing (Smith, 2013; Wilson, 2008).

Participatory observation field notes were unstructured. During the first year of connecting with community members and during data collection, field notes detailed the name, date, and duration of event, and information on community members. Context of the event or interactions were noted, along with my positionality as a non-Native person, as a researcher, as a fellow person of color, and as a woman who experienced violence. Any learned cultural norms, processes, and terms were also noted. A separate document of reflections or memos were kept that detailed internal reflections of the my thoughts, emotions, questions, and connections made with other events [26].

## Ethics statement

The study was approved by Oregon State University’s Institutional Review Board (IRB-2020-0671). Verbal consent was obtained from all participants.

### Relational accountability

As a non-Indigenous scholar, I practiced reflexivity throughout the research process [27]. This involved keeping a journal of expectations, reflections of values, and questions. In addition, I frequently met with partnered community leaders to discuss method development, results, and dissemination of findings. My positionality as a non-Indigenous academic researcher interacting with Native community members of academic and non-academic backgrounds was constantly examined to minimize reproducing colonial dynamics of harm imposed by academic institutions on Indigenous communities [28]. Discussions with community members often did not involve preconceived intentions of the research. Rather, community members were asked about their perspectives on issues of violence against AIAN women and what they perceived to be a need to prevent it.

Community leaders at the Indigenous Circle of Wellness and Seven Generations Child and Family Services supported and approved of the research direction. Principles of non-intrusive observation, deep listening with more than the ears, reflective non-judgmental consideration, etc. were incorporated to ensure ethical responsibility and sensitivity [19]. The research direction and design were developed over multiple meetings in which community leaders emphasized the importance of creating a space that was Native centered in all stages of the research. For instance, “support-seeking” was used in place of “help-seeking” to reduce paternalistic assumptions that AIAN women need “help,” and highlight the agency of survivors in their decisions to seek support. Recruitment flyers and terminologies were examined for familiarity and relatability among Native communities. Instead of using “focus groups,” “Community Circles” was used as it was culturally appropriate and familiar to talking circles [19]. It was also a culturally appropriate way to seek input and enabled me to “view from below” rather than accentuating my positionality as an academic extracting information from a sub-group of people who are subject to the gaze of a researcher [23,29]. Further, a community leader from either partner organization, who are also mental health therapists, always attended a Community Circle session, increasing trust among community members and allowing me to be accepted and welcomed into the space.

### Community Circles and Oral History

Community Circles were formatted as talking circles for Indigenous women to gather and have an equal chance to speak and be heard. The general goal of Community Circles was for women to engage in the relational healing [30] and share their experiences on seeking support as survivors of IPV. This is different from focus group discussions, in which the researcher is looked upon as the main facilitator of the direction of the discussion. Rather, Community Circles as a familiar space enabled the participants to actively engage in dis NN cussing their experiences and easily apply talking circle processes of listening, sharing, and uplifting. I was introduced by the trusted community leader to the participants and a community member would volunteer to open with a prayer. An overview of the project was presented along with a gift choice of a beading kit from a local Native artist or a $25 gift card. At the end of the Community Circle, participants had the option of scheduling a separate one-on-one session with me to share their Oral Histories. Every Community Circle was attended by a professional Native mental health therapist as support.

Similar to Community Circles, Oral Histories were unstructured with the general goal of survivors sharing their experiences of seeking support. It was a dedicated space for AIAN women to tell their own stories. Oral Histories were conducted with Community Circle respondents due to me having established a relationship with the participant from a Community Circle session. This allowed for a trusted setting in which the participant shared details on their family history, childhood, healing processes, and aspirations for the future. Individual nuances of how they navigated various events in their life contributed to the larger understanding of why and how they sought support as survivors. Oral Histories are part of many Indigenous traditions in which language, culture, and relationships are preserved. However, each tribe has their unique language, identity, and cultural practices that cannot be essentialized. Oral Histories were used to emphasize the nuanced experiences of AIAN women survivors who come from different tribes, experiences, and family histories [31]. Participants in this study are from the following tribes: Bishop Paiute, Cahuilla, Chickasaw, Choctaw, Chumash, Comanche, Diné (Navajo), Ho Chunk, Mashpee Wampanoag, Menominee, Mvskoke, Nahua, Pechanga, Pueblo, Seminole, Seneca, Taos, and Tongva. Not all participants knew their tribal affiliations. This study provides a snapshot from the voices of those who shared their stories and cannot be essentialized.

### Sampling

Four Community Circles and 10 Oral History interviews were completed. Community Circle participants were recruited via convenience sampling. Flyers for recruitment were distributed via the Indigenous Circle of Wellness and Seven Generations Child and Family Services’ e-mail newsletter and on social media (e.g., Instagram, Facebook, and Twitter). Interested individuals were led to a web link to determine whether they met the eligibility criteria and confirm that they had access to a safe environment to participate. Eligibility criteria included individuals who:

- 18 to 64 years of age
- Self-identified as Indigenous and/or Alaska Native/American Indian
- Self-identified as a woman
- Experienced violence from an intimate partner

Community Circles were 90 to 120 minutes with four to eight participants and were conducted via Zoom due to COVID-19 safety restrictions. Participants were given the option to leave their camera off and contribute via the Zoom chat box. Several participants did interact with the group in this manner due to their children being present or because they felt uncomfortable showing their faces in a group setting.

Community Circles began with a community leader introducing me to the group. I presented a short introduction about myself and the research project. IPV was clearly defined to make explicit that multiple forms of violence can also be considered IPV. Participants were informed that they were not required to share details of their IPV experience. They were also provided with clear communication pathways to me, such as e-mail and phone. Updates on the progress of the research were sent out after each Community Circle and during analysis. Verbal consent was obtained from all participants and the session was audio recorded. After each Community Circle session, an invitation to participate in an Oral History interview and a list of local and national resources were sent. Feedback was also solicited in the post-session e-mail and debriefs with the community leader present for the session were done immediately after the Community Circle ended.

Demographic information from Community Circle participants was not gathered to minimize replicating academic research settings and allow the participants to reveal as much information about themselves as they desired. To protect their confidentiality, their names and tribal affiliations will not be identified in the results.

A total of 10 Oral History interviews were conducted, nine of whom were recruited from Community Circles and one who was unable to attend a Community Circle but volunteered to share their Oral History. Oral Histories were semi-structured, focusing on the participant’s background, their family history, and more details on their support-seeking decisions. Oral histories were two hours in length and conducted via Zoom due to COVID-19 restrictions. Participants were able to choose another $25 gift in the form of a gift card or beading kit. Once verbal consent was obtained from the participant, the session was audio recorded.

Demographic information was collected in this setting as part of understanding the individual’s oral history narrative. Table 1 provides a demographic summary of Oral History participants (n=10). Participants resided in both urban and reservations in California. Participant ages ranged from 30 to 51. Half of the participants identified as heterosexual, 20% as bisexual, 10% as queer, and 10% as pansexual. For formal education, 60% of the participants obtained or were obtaining a graduate degree. Forty percent attended some college or trade school. Eighty percent of participants were employed at the time. As for marital status, 60% identified as single, 10% as married, 20% as separated or divorced, and 10% as co-habiting. Finally, 30% of the participants identified their ethnic background as Indigenous only and the remaining 70% identified as Indigenous and as other ethnicities.

**Table 1.** Demographic characteristics of Oral History participants

| Demographic characteristics | Participants (n=10) |
| --- | --- |
| Age (average) | 37.3 |
| *Sexual Orientation (%)* |  |
| Heterosexual | 50 |
| Queer | 10 |
| Bisexual | 20 |
| Pansexual | 10 |
| No Answer | 10 |
| *Education (%)* |  |
| Some college or trade school | 40 |
| Bachelors | 0 |
| Graduate | 60 |
| Employed (%) | 80 |
| *Marital Status (%)* |  |
| Married | 10 |
| Single | 60 |
| Separated or Divorced | 20 |
| Co-habiting | 10 |
| *Ethnic background (%)* |  |
| Indigenous only | 30 |
| Multi-ethnic | 70 |

Participants were notified that they could request transcripts of their Oral Histories. Two requested transcripts but there was no further feedback provided once the transcripts were received. At the conclusion of Community Circle and Oral History interviews, a community report with initial findings was distributed to all participants for review. The Indigenous Circle of Wellness and Seven Generations Child and Family Services approved the community report and three participants who engaged in a Community Circle and Oral History responded with positive feedback.

## Analysis

Audio recordings were transcribed using Otter.ai transcription and transcripts were analyzed using MAXQDA software. All transcripts underwent initial *in vivo* coding to ground the analysis in the survivors’ perspectives. Concepts that emerged from the *in vivo* coding process informed our results. Community Circle transcripts were analyzed separately from Oral Histories to observe whether similar codes and themes would emerge. Initial findings from the first round of coding and thematic analysis were compiled into a community report that was distributed to the community partners and participants for review. The second round of coding was completed based on community feedback. Participants resonated with stories of healing and the collective need for Native-only community spaces for survivors. Thus, the second round of coding revisited *in vivo* codes that included topics of healing and community and delineated how healing and community were characterized.

## Reliability

As a non-Indigenous researcher representing an academic institution, every decision and interaction were done with intention and accountability. Once I was connected to key community leaders at Indigenous Circle of Wellness and Seven Generations Child and Family Services, I sent a research brief that detailed who I was beyond my academic identity. During initial meetings with community partners, I was asked what my connection was to California as I did not reside there at the time of this research and writing. We connected on my family’s history as immigrants who first lived in California, and my childhood memories of traveling to California frequently to access Koreatown and have a semblance of home. As a first-generation US citizen, I grew up with a multicultural identity. This story carried on into Community Circles as I introduced the study to the participants. As an outsider, the women were interested in my motivations and my connection to Native people (Smith, 2013). This project relied heavily on the community partners’ feedback, and I frequently met with the community leaders from the Indigenous Circle of Wellness and Seven Generations Child and Family Services to reflect on Community Circles and implement any changes suggested by the participants. Member checking was prominent throughout the analysis phase to ensure that the developing themes reflected the voices and perspectives of the participants.

## Results

Women used multiple strategies to seek support. Formal networks of support include therapy with a mental health professional, safe houses, medical care, housing services, community advocacy services, and law enforcement. Informal networks of support include loved ones, such as friends, family, and partners. While past studies have explored predictors of seeking support, we are further interested in the dynamic nature of seeking support over time, and the relationships between the different areas of support and with the survivor. Survivor stories in Community Circles and Oral History interviews reveal four salient relationships: 1) Informal support is entangled with intergenerational trauma and normalization within families; 2) Some forms of formal support can be harmful to the survivor, therefore solidifying Indigenous survivors’ mistrust in these institutions; and 3) Healing is found in ceremony; and 4) Support that is community-based and culturally informed is imperative for Indigenous survivors.

### Entanglements with informal support

In many cases, friends and family are the first line of support that survivors seek out. As Native women, seeking support from family or their Indigenous community is complex. The ripple effects of colonization and genocide of Native peoples, and the subsequent intergenerational trauma that occurs is often a focal point when survivors reflect on their histories of abuse. Thus, when family members respond by victim-blaming or normalizing the abusive relationship, this response is attributed to the normalization of family violence and intergenerational trauma that their families have experienced.

> And I had moved into my grandmother’s house to tell my Grandma what was happening. You know, what he was doing to me. And after I told her, she told me, well, “you made your bed now lay in it.” (Community Circle 1)

Nods of agreement or typed affirmations in the Zoom chat box followed as this survivor recalled her grandmother’s response. One community member wrote, “*my mom said the same thing*,” further reflecting the depth in which the victim-blaming and normalization of IPV have passed through generations.

> When you grow up, and there’s a lot of sexual and physical violence present, and you’re like, these hurt. This shouldn’t be happening. And then people are telling you, “we do this anyway.” You get such a distorted vision of the world and, and lots of times when I’m not feeling myself, I’m in my head spinning around in there…I don’t have any kind of clarity. (Community Circle 1)

Survivors will often reflect on their family and friends’ responses that have fallen short. In hindsight, many can rationalize reasons for why they were met with such reactions. For example, when one survivor was asked if she reached out to friends for support, she describes how her friends’ lack of support could be attributed to the lack of information and education available then on how to support survivors.

> They weren’t dismissive, but it was, I suppose because they didn’t know what to do. This was like in the late 90s, early 2000s. So there really wasn’t this discussion of like, how to help survivors or what to do if someone comes to you. And this was like, you know, the internet just came about. (Oral History 4)

Similar rationalizations are expressed regarding family members who have also responded to survivors in non-supportive ways. Particularly for family members that victim-blamed the survivor, the women recall the experience with an understanding of why their loved ones responded this way. Survivors acknowledge the complexity of trying to find healing within the very family units that have experienced generations of systemic violence and have internalized victim-blaming behavior.

> Just thinking Indigenous wise, and tribal wise, [it] really does take a village to raise anybody…I believe in that and I believe in the family because I’ve gotten so much support and healing from my family, I really have. But it’s hard because it’s kind of like trying to fix something with the thing that’s broken. Like seeking support from the people who installed my buttons… it’s hard to heal within that family unit. (Oral History 8)

All the women in this study describe the role that violence played throughout their family as they recount generations of women who have also endured violent relationships. They often attribute the existing history of violence within their family in relationship to traumatic policies like being placed in boarding schools, the Dawes Act, etc.

> Now, my grandmother was raised in boarding schools since she was three. She stayed there until she was 18. So, I believe that, even though she said that at that time, I took it to heart at that time. But now I realize that it was we talk about multi-generational trauma, that she didn’t grow up in a loving family or even having the ability to show love or give love because it was never given to her. (Community Circle 1)

While some family members, particularly mothers and grandmothers, may normalize IPV, the survivors themselves express a refusal to allow this “generational curse” to continue. This resolve is borne from more supportive interactions with loved ones. Friends, family, or partners can offer strong support to survivors as they navigate leaving their abusers and begin the recovery or healing process.

For one participant, an extended family member was crucial to her acknowledging the IPV in her relationship at the time. What distinguished this family member from others was their ability to name the abuse and “just [be] honest, just listening… [they weren’t] there to judge.”

> I think that’s when [family member] was a big thing because he never just told me to like leave or anything like that…He was just like, “it’s domestic violence. I care about you.” So, he’d take me out to dinner every now and then just to check up on me. And it wasn’t frustration. It was more like, “I just want to make sure you’re okay.” (Oral History 2)

This survivor identifies her family member’s support as being exemplary of love and patience that clarified the type of support the survivor needed. She mirrors the same kind of support she received from her family member to a friend experiencing IPV by extending grace and patience as opposed to pushing them to leave the relationship.

Another survivor describes her parents as being “bodyguards” as they directly intervened when her abuser tried to reach her. In one instance, while she tried to break up with her abuser, he forced himself into her home. The survivor called her mother, and the abuser took the phone from her. He claimed that she was suicidal and that he was going to take her to the emergency room to get checked in.

> My mom was like, super cool, she’s like “Oh, thanks for telling me. You don’t need to take her to the hospital, I’ll come home, I’ll take her right now.” Superconscious. “Give the phone to [survivor].” So, he hands me the phone, she’s not on speaker anything. And she was like, “try and stay like in public. Don’t be in the house alone with him. I’m coming home right now,” and she just knew. She totally knew. (Oral History 3)

Her father also served as a shield by refusing to let the abuser contact her. Her parents never questioned the validity of the abuse and were quick to protect their daughter from further interactions or potential harm from the abuser.

Such examples of seemingly unconditional support from loved ones for survivors were rare among the participants in this study. While we cannot postulate that this is representative across all Native women survivors, we observe patterns of support from loved ones, especially family, is intertwined with intergenerational trauma and violence.

### Inadequate outsider systems

When support from loved ones is rife with victim-blaming or lack of acknowledgment, survivors have accessed other forms of support. Formal networks were often mentioned as emergent sources of support due to feeling endangered or because the survivor needed immediate attention from a violent interaction.

Law enforcement is especially regarded among the survivors as an untrustworthy source. Some have contemplated reporting or seeking a restraining order, but their mental map of what would accumulate often leads to exhaustion and doubt that police will be a beneficial resource.

> I really wanted to get a restraining order, but it was so much exhaustion of what I was doing where I didn’t want to get that restraining order because I have to talk about you know, everything he does to me mentally and then also physically and I had-something I didn’t want to really relive. And it’s something that I don’t think police, men, or even the police system, in general, would believe me. (Community Circle 1)

Multiple women in the study recall moments where they were put in handcuffs or blamed by judicial authorities. Across all the women in this study, none sought out police willingly as a primary source of support. Rather, their interactions with the police were often a result of the abuser using law enforcement as a tool to control or coerce the women.

> I never have trusted the cops ever. My whole life I’ve never trusted the police so that was definitely not a safe place. Oddly enough, he called the police on me one time when I was telling him that I was gonna leave. He called the cops and told them that I was having a psychotic break and I had threatened him, so they came into my home, put handcuffs on me. (Oral History 9)

For some women, even when they are no longer in an intimate relationship with the abuser, the abuser continues to manipulate or harass them. Survivors describe how ex-partners will use their children as pawns to harass and abuse the survivor. In other cases, ex-partners will continue to stalk the survivor. Such chronic exposure to violence from an intimate partner unfairly burdens the survivor to then manage their safety and boundaries, all the while they are trying to heal from the traumatic experience.

> And it’s a way that he kind of still controls me to this day and, and I want to delete all of like, I mean, I have fucking hundreds of pages of just, “He didn’t show up at this time. Then he came two days later and he screamed and yelled about this and then did just all this stuff.” And it’s so crazy-making, like they really asked me to just like sit in it, to be so fully, fully consumed with that relationship. How the hell are you supposed to heal or like move on to anything else? (Oral History 5).

Another survivor who is also a mother recalls how her ex-partner also uses law enforcement and judicial courts to his advantage. In their disputes regarding their children, the abuser will continue to take her to a court or even call the police on her to obtain his objective. One incident involved the abuser calling the police on the mother while their children were staying with her.

> I had the kids [and] he thought he should have them. But it wasn’t his time. And I told him not to come, that I wasn’t gonna release the children… I was within my rights. And he came anyway and knocked on my door. And I was like, “I’m not releasing the children.” Then he brought the police. And there was nothing the police could do either.

When the police came, the way they appeared gave her the impression that they saw her as the perpetrator.

> …they came in two separate police cars and they were both wearing flak jackets. I was thinking he must have told them the kids are in danger or something like very serious…they came like within five minutes…They don’t normally wear flak jackets around [here] and [he] must have portrayed it as some kind of dangerous domestic dispute.

As she reflected on this incident, she continued to discuss how her ex-partner’s decision to call the police reflects his disregard for their children’s safety. Rather, it was an act of exerting power by mobilizing law enforcement against her.

> And afterward… [my youngest child] was terrified, because the policeman was so big and white and mom was so small and brown, and he was sure the policeman was gonna shoot into the house or shoot me or something. But like, [x] didn’t even consider like, how does this affect my kids, but call the police on my ex-wife, on their mom, who’s there in the house with them? So, I don’t think he even thinks about those kinds of things. (Oral History 6)

Just as many individual Native women express distrust in the police, the relationship between Native communities and law enforcement are entangled in the survivor’s decision to call the police. For example, one woman shared how her reservation has an unspoken rule to not call the police. However, her decision to call ultimately stemmed from the need to have something on record to benefit her case for custody of her child in the future.

> So when the police came looking for him, and they were questioning people, there were women who actually called my husband at the time and told him that, “you better look out because the police came looking for you,” and everything like that. He could have killed me… It was a terrible situation…I wish there was somebody else I could have called besides the police to help me out, you know, I wish that there was something else that I could have done. (Community Circle 1)

Inherent mistrust from the community entangled with the need to find protection for the family led to the reservation community and the outside legal system harming the survivor. This survivor’s relationship with her community positioned her at the crosshairs of breaking her community’s “rule” versus prioritizing her and her child’s safety by seeking the only legal source available. None of the women in this study recall their relationship to law enforcement and judicial services as supportive. Rather, while it symbolically may serve as an institution of protection, it is regarded with intense mistrust and unreliability in supporting survivors of IPV.

### Healing is ceremony

In both Community Circles and Oral History interviews, the women were asked about what healing meant to them, what worked, and what they wished was available to fellow Indigenous women survivors of IPV. Survivors in this study primarily sought healing in Indigenous-specific formal services (e.g. therapy) and community. Healing is often described in settings of the ceremony, such as engaging in an intimate process with the tribal community:

> “You spend a year preparing… the initial ceremony where you go to ask for help…You have to bring offerings, you have to bring food, you have to like, do all these protocols, but then you also have to speak and tell the medicine man your reasons.” (Oral History 6)

Ceremony with tribal elders is described as a way for survivors to reconnect with their roots:

And so when I go see elders and I do the ceremony, or I do different things, it helps me kind of find like my roots again so that I’m not just spinning around. (Oral History 5)

Survivors have also engaged in ceremonies alone. Particularly for those that have been estranged from their tribes or Indigenous history, reconnecting with their ancestors’ traditions was foundational to their healing process. One participant describes using salvia, a sacred plant used by her people in ceremony “that calls your heart back into place.”

So I went through the ceremony. And I actually felt my ancestors, like standing beside me, like I felt their hands like on my shoulder, and it was a particularly, just healing and moving experience. I felt the beat, like their heartbeat, and our hearts just beat as one…for the first time in my life. I felt whole, I felt at peace. And so, being given that medicine was just a particularly healing and life-changing moment. (Oral History 4)

Beading was frequently mentioned in both Community Circles and Oral History interviews as integral to their healing. Beading serves a meaningful purpose as it connects Natives to their culture and history, and are used to create or decorate jewelry, clothing, or regalia. Beadwork tells stories and is intimately valued [32]. For one survivor, she describes how people encourage her to sell her beadwork, but she does not see it for its monetary value: Any type of beading is medicine… It’s what I have overcome, I mean beading has helped me…it’s my medicine. (Community Circle 4)

It is also a vehicle for collective bonding that Native people can engage in. While each tribe may have its own significant meanings behind their beadwork, tribes like the Choctaw, Seneca, Chickasaw, Tongva, Cherokee, Diné, etc. share this spiritual tradition. Thus, beading with other Native members in the community can serve as a healing space.

> …you go into that circle, you’re all working on your own projects, it could be completely silent. And it’s just nice to sit there with other people and nobody’s like hashing out these big life secrets, or, like these intense traumas, but it’s just that it’s like an extended family, you know, that’s just super supportive. And it’s all there with love in our hearts for everybody. (Oral History 3)

Due to most of the participants having been recruited through community partners that provide mental health services, it was unsurprising to learn that most of the women had sought or are seeking therapy. In many ways, therapy or support groups were crucial to their healing journeys, especially if the therapist was Indigenous. One survivor listed the identities of her therapist that were particularly impactful for her:

> Yeah, so fit for me was one: she identified as Native. That was really huge for me. And not only Native, but also Xicana as well, because I come from that household, you know, my dad is Mexican. And then being just also Native was really important. And being a woman was really important. (Oral History 2)

It further served as an avenue for survivors to come to recognize and label the various acts of violence they experienced throughout their life. Particularly for those who witness violence throughout their upbringing, and it was normalized, therapy was pivotal to shifting their understandings of what violence could look like.

> But going to therapy was a really big eye-opener to recognize the kind of pathway that I went by accepting violence in my life. (Oral History 2)

Therapy, when culturally centered on Indigenous identity, carved a space for survivors to not only connect to their roots but process their traumas in a way that they could understand from their Indigenous world views. When asked about the difference between Indigenous and non-Indigenous support groups, one survivor shares:

> I feel that in an indigenous way, it’s more personal, more connected to the creator. And I think that’s why I had to go more indigenous because there are more teachings on… creator and how things work together in a full circle.” (Oral History 10)

It also has its mental health benefits as survivors work through their traumas, they feel more empowered to connect deeper with their communities and branch out to other forms of support to aid in their healing processes. For instance, one survivor shared during her Community Circle session that once she was able to find the strength to do therapy and yoga, she was “able to rebuild her community through reconnecting to other Indigenous people.” Broadening her connections and methods of support catalyzed her ability to rebuild “little pockets of communities.” (Community Circle 3)

### Culturally competent and community-based systems of support

Survivors in both Community Circles and Oral History interviews were asked what was needed for Indigenous women survivors and community members. Regardless of tribal affiliations, age, or occupation, they all expressed the urgent need for culturally based services that catered specifically to Indigenous women.

> And then participating in culturally based interventions strengthen our connections. So, for us as Native people, the culture and then coming together like this is an important part of our culture, right, is that connection with each other. So, it’s just great to hear, feeling as well, and you know making sure that it’s not passed on from generation to generation. (Community Circle 4)
>
> But also too, while engaging in, in like, interaction, like, even if it’s a group setting or therapy setting, to have more my cultural elements there. Because as I know that, you know, domestic violence isn’t traditional, yet what I talked about multi-generational trauma, it exists, it’s there. (Community Circle 1)

To be able to access formal services, such as therapy, in settings where there is immediate cultural connection strengthens community ties. Further, being able to engage in environments where identities and traumas do not have to be elaborately explained may provide survivors some respite, opening space to focus on fostering a connection to their traditions and healing.

For Native women who are estranged from their tribal history, seeking connection to their roots is often painful and isolating. One woman recalls how she desperately cried out to her ancestors:

> “I don’t even know exactly who you are, I really need your help.” And I would just go and sit in nature and sit by a tree and cry and that was how I got through it. (Oral History 9)

Investing in culturally competent and community-based services allows for women to reclaim their bodily sovereignty [22]. Beyond healing from the trauma at physical, emotional, and psychological levels, healing that is culturally centered allows Indigenous women to pursue healing as symbolizing resistance to the societal level of historical oppression and resulting intergenerational violence.

> For Native women to have the power and the strength, and the support, we need to be able to reclaim our sovereignty over our spirits and our bodies and the same way that many of us advocate for reclaiming the sovereignty of our tribes. (Oral History 5)

The ability for survivors to reclaim their sovereignty for themselves has a direct impact on their relationships. Among the participants who are mothers, they are determined to reclaim their traditions by breaking the cycle of violence for the sake of their children.

> Like we don’t, you know, they say, like one of the Native women’s orgs that I work with say, you know, violence against women is not traditional. You know, and there may be people, in our communities, in our Native communities that tell us it is, you know, or that would like us to believe otherwise. (Oral History 5)
>
> I want to stop by that cycle, and it has to start with me…I believe it’s a generational curse and until somebody stands up and stops it, it will continue. (Community Circle 4).

To seek support and have access to culturally rooted support systems have significance that impacts beyond the individual level. Rather, survivors invoke the impact that their healing will have on their relatives and community, breaking the cycle of violence for future generations.

## Discussion

Survivors have dynamic journeys as they pursue healing from their IPV experiences. While their paths to finding support may look different from person to person there are apparent commonalities among all women who participated in the Community Circles and Oral Histories. The first two themes discuss the complex relationships between survivors and informal and formal support systems. The latter two themes highlight how healing and support is actualized or imagined by the survivors. Among the Community Circles and Oral History participants, seeking informal support from a friend or family member is frequently mentioned. While this was an expected outcome [11–13], this study allowed for a deeper understanding of how informal support manifested throughout the survivors’ lives. Participants who are younger or experienced violence more recently discussed how social media posts validated their experiences and reminded to seek out resources when needed. Normalization and victim-blaming behavior is observed in families, often among women whose mothers or grandmothers also experienced IPV in their lifetimes. On the other hand, when family members are supportive and validate the experience, the survivors credit such responses to their decision to seek formal support (e.g. therapy) later on [14].

All participants in the study regard law enforcement and the legal system as untrustworthy sources of support. Seeking support from the police is often due to increased risk of violence [11,16,33] and/or to protect their children from the abuser [34]. Police interactions with survivors reveal victim-blaming behavior as police will handcuff the survivor or further the trauma by requiring the women to recall in detail the violent incidents. The legal system is an additional stressor as abusers wield it to control and manipulate the women even after separation. While abusers continue to utilize law enforcement and the legal system in their favor, the women grapple with how to heal from the relationship and find safety for themselves and their children. The need to continue to record all abusive interactions while actively protecting their children result in feelings of exhaustion, anger, and frustration. For one woman in Community Circle 1 who shared that the women in her community warned her abuser about the police coming is exemplary of a tight-knit community that can act as a barrier to seeking formal support. This woman’s decision to call the police was not primarily motivated by trust, but the need to have something on record to fight for custody of her child. However, her tight-knit community setting’s deep mistrust in the police exacerbated the woman’s risk of experiencing violence. Mistrust in the overall legal system permeates the institutional, community, and individual levels as women weigh their decision to call the police or go to court.

Informal and formal support systems can be harmful and inadequate in meeting the survivors needs. When asked about how they pursue healing and what a supportive system looks like, ceremony and culturally rooted systems of support are frequently mentioned. To engage in healing as ceremony is unique to Indigenous women as it is culturally specific and an unmet need among Native women survivors. As a population that has significantly high risk of experiencing IPV and are significantly more likely to lack access to needed services than non-Hispanic White women (Rosay, 2016), healing from IPV is an arduous and, sometimes, a self-guided process. Ceremony with their relatives and tribal members can be exemplary of a supportive tight-knit community in which survivors can engage in ceremony with a community healer or through beading circles. Communal healing spaces that are safe and specific to Native women allows the women to connect with each other not only as survivors of IPV, but as fellow Indigenous women, as mothers, as daughters, and as extended relatives. For some women who participated in the Community Circles, this was the first time they had been in a protected space only for Native women survivors. After the Community Circles ended, participants asked when they could participate in the next one and expressed their desire to engage in more spaces like the Community Circle as it was particularly healing to share and collectively bond with other survivors. One woman described how it felt “effortless” by being able to show up and not feel burdened to explain how IPV is intertwined with her Indigenous identity.

Support systems that can transcend beyond the individual and strengthen the community and institutional levels are critical for Native women survivors. Continued education and outreach to individuals and community members on IPV is important but having information that is specific to Native people is imperative to fighting public health’s history of pushing AIANs to the margins of public health research and interest. Community Circles are exemplary of a protected space that permeates through all levels of the ecosystemic framework. It is a community setting in which individuals can participate in. Each Circle had a professional Indigenous mental health therapist present from one of the partner organizations. Institutional representation that is Indigenous and culturally rooted increases trust among survivors as well. Therefore, support for AIAN women survivors of IPV must be robust in empowering their cultural identities and increasing the availability of community-centered support systems.

## Limitations

COVID-19 restrictions prevented in-person Community Circles and Oral History interviews. This had its benefits as women who may not have had the courage to share their stories in person felt more comfortable attending a Community Circle and participating with their videos off. However, occasional audio and connection challenges did occur.

Most participants discuss seeking mental health support as part of their healing journeys. While this was expected due to the nature of recruitment, this may not be representative of the many other Indigenous women across the Americas who have experienced IPV. Among Oral History participants, 60% of the women have graduate degrees. This is not representative of the larger AIAN population. Higher levels of educational attainment is associated with support-seeking behaviors [11,33]. Whereas most of the women in Community Circles do not have graduate degrees, those who further opted in to share their Oral Histories may be partially attributed to their education status. In addition, these women were also familiar with therapy and demonstrated the ability to speak on their experiences more cohesively. Despite their involvement with therapy and their healing journeys, gaps in adequate and culturally rooted support was evident. This may indicate that we are underestimating the impact of IPV and need for safe healing spaces for those who do not have a graduate degree or are not connected to Indigenous institutions like Indigenous Circle of Wellness and Seven Generations Child and Family Services. The community partners are in urban California cities and the experiences of the participants in this study do not wholly represent the experiences of those who have lived or currently live on reservations.

The method of this research utilizes a more participatory approach in which the researcher engages with the community. In doing so, the relationship between the researcher and the participants is unique. The researcher in this study does not identify as Indigenous which may result in limited understandings or incomplete interpretation of data. A collaborative approach and significant support from Native community leaders was critical to ensure relational accountability and rigor of the research.

Possible future research includes doing a closer follow-up with the participants to garner feedback on study processes and findings, or increasing feedback received when sharing transcripts and results with the participants. Women in this study often connected their Native identities and tribal histories to their experiences of seeking support. Harmful governmental policies like the Dawes Act and previous generations’ survivorship from boarding schools have shaped the family’s understanding of violence. The impact of such policies aiding and perpetuating dysfunctional family perceptions on IPV needs to be further pursued. Lastly, targeting recruitment to include not only those who identified as women but also as two-spirit introduces diversity of identities and experiences as survivors of IPV.

## Conclusion

Violence against Indigenous women is an issue that affects survivors physically, mentally, emotionally, and spiritually. Intertwined with histories of oppression and colonization, historical oppression is often an essential component to understanding IPV among AIANs [5,35– 37]. The concept of “soul wounds” is an Indigenous paradigm for various physical, mental, and emotional effects of violence. For AIAN women, experiencing and healing from violence is a laborious process as the resulting trauma can have intergenerational effects that perpetuate a cycle of violence within their families [38].

Navigating a world where there is a persistent lack of funding for culturally competent services, fear of medical and legal personnel not being sympathetic towards Natives due to misperceptions, stereotypes, and prejudice are all factors that make combating the issue of violence against Native women even more difficult [37]. Seeking support and healing for Native women survivors of IPV is layered with their identities as Indigenous women, historical oppression, and its resulting intergenerational traumas. Healing from the violence has cultural implications, amplifying the need for community-based systems of support that are Indigenous-centered. For AIAN women survivors of IPV, healing transcends beyond their selves, extending to their families, tribal communities, and future generations. As one participant described: *I am the newest branch of a tree that is ancient*. (Oral History 3)

## Data Availability

Participants were recruited through direct partnership with the Indigenous Circle of Wellness and Seven Generations Child & Family Services in California and can be easily identified. Therefore, the data cannot be ethically shared.

## Acknowledgements

My deepest gratitude to the women who shared their stories so powerfully. I thank Monique Castro, LMFT from the Indigenous Circle of Wellness and Dr. Carrie Johnson from Seven Generations Child & Family Services for their support.

## Notes

### Competing Interest Statement

The authors have declared no competing interest.

### Funding Statement

Melissa Institute for Violence Prevention and Treatment The funder had no role in study design, data collection and analysis, decision to publish, or preparation of the manuscript.

### Author Declarations

The study was approved by Oregon State University's Institutional Review Board (IRB-2020-0671). Verbal consent was obtained from all participants. Oregon State University A312 Kerr Administration Corvallis, OR 97331-2140 Phone 541-737-3467 Fax 541-737-9041

## References

1. Echo-Hawk A. Our Bodies, Our Stories: Sexual Violence Among Native Women in Seattle, WA [Internet]. Urban Indian Health Institute; 2018. Available from: http://www.uihi.org/wp-content/uploads/2018/08/UIHI_sexual-violence_r601_pagesFINAL.pdf

2. Evans-Campbell T, Lindhorst T, Huang B, Walters KL. Interpersonal Violence in the Lives of Urban American Indian and Alaska Native Women: Implications for Health, Mental Health, and Help-Seeking. Am J Public Health. 2006 Aug;96(8):1416–22.

3. Tjaden P, Thoennes N. Full Report of the Prevalence, Incidence, and Consequences of Violence Against Women: (514172006-001) [Internet]. American Psychological Association; 2000 [cited 2020 Jan 17]. Available from: http://doi.apa.org/get-pe-doi.cfm?doi=10.1037/e514172006-001

4. Wahab S, Olson L. Intimate Partner Violence and Sexual Assault in Native American Communities. Trauma Violence Abuse. 2004 Oct 1;5(4):353–66.

5. Walters KL, Simoni JM. Reconceptualizing Native Women’s Health: An “Indigenist” Stress-Coping Model. Am J Public Health. 2002 Apr;92(4):520–4.

6. Bachman R, Zaykowski H, Lanier C, Poteyeva M, Kallmyer R. Estimating the Magnitude of Rape and Sexual Assault Against American Indian and Alaska Native (AIAN) Women. Aust N Z J Criminol. 2010 Aug 1;43(2):199–222.

7. Deer S. The beginning and end of rape: confronting sexual violence in native America. Minneapolis: University of Minnesota Press; 2015. 207 p.

8. Rosay AB. Violence Against American Indian and Alaska Native Women and Men. 2016 Sep;8.

9. Walters KL, Simoni JM. Decolonizing Strategies for Mentoring American Indians and Alaska Natives in HIV and Mental Health Research. Am J Public Health. 2009 Apr 1;99(S1):S71–6.

10. CDC. Preventing Intimate Partner Violence [Internet]. CDC. 2019 [cited 2019 Dec 7]. Available from: https://www.cdc.gov/violenceprevention/intimatepartnerviolence/fastfact.html

11. Goodson A, Hayes BE. Help-Seeking Behaviors of Intimate Partner Violence Victims: A Cross-National Analysis in Developing Nations. J Interpers Violence. 2018 Aug 23;0886260518794508.

12. Gover AR, Tomsich EA, Richards TN. Victimization and Help-Seeking Among Survivors of Intimate Partner Violence. 2015 Mar 4 [cited 2019 Nov 22]; Available from: https://www.oxfordhandbooks.com/view/10.1093/oxfordhb/9780199935383.001.0001/oxfordhb-9780199935383-e-58

13. Kim E, Hogge I. Intimate Partner Violence among Asian Indian Women in the United States: Recognition of Abuse and Help-Seeking Attitudes. Int J Ment Health. 2015 Jul 3;44(3):200–14.

14. Evans MA, Feder GS. Help-seeking amongst women survivors of domestic violence: a qualitative study of pathways towards formal and informal support. Health Expect. 2016;19(1):62–73.

15. Burnette CE. Family and cultural protective factors as the bedrock of resilience and growth for Indigenous women who have experienced violence. J Fam Soc Work. 2018 Jan 1;21(1):45–62.

16. Spencer RA, Shahrouri M, Halasa L, Khalaf I, Clark CJ. Women’s Help Seeking for Intimate Partner Violence in Jordan. Health Care Women Int. 2014 Apr 3;35(4):380–99.

17. Burnette CE, Figley CR. Historical Oppression, Resilience, and Transcendence: Can a Holistic Framework Help Explain Violence Experienced by Indigenous People? Soc Work. 2017 Jan 1;62(1):37–44.

18. Odom SK, Jackson P, Derauf D, Inada MK, Aoki AH. Pilinahä: An Indigenous Framework for Health. Curr Dev Nutr. 2019 Feb 22;3(Suppl 2):32–8.

19. Wilson S. Research is Ceremony: Indigenous Research Methods. Black Point, N.S: Fernwood Pub; 2008. 123 p.

20. Duran E, Duran B. Native American postcolonial psychology. Albany: State University of New York Press; 1995. 227 p. (SUNY series in transpersonal and humanistic psychology).

21. Mookerjee S, Cerulli C, Fernandez ID, Chin NP. Do Hispanic and Non-Hispanic Women Survivors of Intimate Partner Violence Differ in Regards to Their Help-Seeking? A Qualitative Study. J Fam Violence. 2015 Oct 1;30(7):839–51.

22. Deer S. Sovereignty of the Soul: Exploring the Intersection of Rape Law Reform and Federal Indian Law. SUFFOLK Univ LAW Rev. 2005;12.

23. Skeggs B. Feminist Ethnography. In: Handbook of Ethnography [Internet]. 1 Oliver’s Yard, 55 City Road, London England EC1Y 1SP United Kingdom: SAGE Publications Ltd; 2001 [cited 2022 Feb 18]. p. 426–42. Available from: http://methods.sagepub.com/book/handbook-of-ethnography/n29.xml

24. Bhattacharya K. Fundamentals of qualitative research: a practical guide. New York: Routledge, Taylor & Francis Group; 2017. 204 p.

25. Burnette CE. From the Ground Up: Indigenous Women’s after Violence Experiences with the Formal Service System in the United States. Br J Soc Work. 2015 Jul 1;45(5):1526–45.

26. Schubotz D. Participatory Research: Why and How to Involve People in Research [Internet]. 1 Oliver’s Yard, 55 City Road London EC1Y 1SP: SAGE Publications Ltd; 2020 [cited 2022 Feb 19]. Available from: https://methods.sagepub.com/book/participatory-research

27. Aronowitz R, Deener A, Keene D, Schnittker J, Tach L. Cultural Reflexivity in Health Research and Practice. Am J Public Health. 2015 Jul;105(Suppl 3):S403–8.

28. Smith LT. Decolonizing methodologies: research and indigenous peoples. Second edition. London & New: Zed Books; 2012. 240 p.

29. hooks bell. Talking back: thinking feminist, thinking black. Boston, MA: South End Press; 1989. 184 p.

30. Mehl-Madrona L. Healing relational trauma through relational means: aboriginal approaches. In: Potter N, editor. Trauma, Truth and Reconciliation [Internet]. Oxford University Press; 2006 [cited 2022 Feb 1]. p. 277–98. Available from: http://oxfordmedicine.com/view/10.1093/med/9780198569435.001.0001/med-9780198569435-chapter-012

31. Mahuika N. Rethinking Oral History and Tradition: An Indigenous Perspective [Internet]. 1st ed. Oxford University Press; 2019 [cited 2022 Feb 19]. Available from: https://oxford.universitypressscholarship.com/view/10.1093/oso/9780190681685.001.0001/oso-9780190681685

32. Gray MJ. Beads: Symbols of Indigenous Cultural Resilience and Value. 2017;51.

33. Tenkorang EY, Sedziafa AP, Owusu AY. Does Type and Severity of Violence Affect the Help-Seeking Behaviors of Victims of Intimate Partner Violence in Nigeria? J Fam Issues. 2017 Oct 1;38(14):2026–46.

34. Rasool S. Help-Seeking After Domestic Violence: The Critical Role of Children. J Interpers Violence. 2016 May 1;31(9):1661–86.

35. Razack SH. Sexualized Violence and Colonialism: Reflections on the Inquiry into Missing and Murdered Indigenous Women. Can J Women Law [Internet]. 2016 Aug 18 [cited 2019 Dec 6]; Available from: https://www.utpjournals.press/doi/abs/10.3138/cjwl.28.2.i

36. Sotero M. A Conceptual Model of Historical Trauma: Implications for Public Health Practice and Research [Internet]. Rochester, NY: Social Science Research Network; 2006 Fall [cited 2019 Dec 11]. Report No.: ID 1350062. Available from: https://papers.ssrn.com/abstract=1350062

37. Weaver HN. The Colonial Context of Violence: Reflections on Violence in the Lives of Native American Women. J Interpers Violence. 2009 Sep 1;24(9):1552–63.

38. Mercy JA, Rosenberg ML, Powell KE, Broome CV, Roper WL. Public health policy for preventing violence. Health Aff Proj Hope. 1993;12(4):7–29.

